# A new approach using proxy event in prior event rate ratio for terminal event studies

**DOI:** 10.64898/2026.06.25.26356521

**Authors:** MA Zhijie, Yong Xiang, SO Hon-Cheong

## Abstract

**Purpose:** This study introduces a novel approach to address unmeasured confounding in terminal event studies using the prior event rate ratio (PERR) method. The proposed approach *PERR_proxy_* used a proxy event to replace the original terminal event in the pre-exposure period, enabling the application of PERR in terminal event settings. Additionally, we also applied difference in difference (DID) regression, which is conceptually analogous to PERR to estimate the standard errors and confidence intervals of *PERR_proxy_*.

**Methods:** We conducted numeric simulations to evaluate the validity of *PERR_proxy_* approach and assessed its performance under varying levels of unmeasured confounding effects, baseline hazard ratios, and the correlation between the proxy and terminal events. To demonstrate its practical applicability, we also performed an empirical analysis to investigate the impact of severe hospitalized COVID-19 on circulatory system disease mortality using the *PERR_proxy_*.

**Results:** In simulation studies, *PERR_proxy_* effectively reduced the unmeasured confounding effects compared to the conventional methods. The performance of *PERR_proxy_* was influenced by the strength of unmeasured confounding, baseline hazard ratios, and the correlation between the proxy and terminal outcomes. In addition, difference in difference (DID) regression had much faster computational speed for estimating standard errors and confidence intervals compared to bootstrap. In the empirical analysis, *PERR_proxy_* identified that severe hospitalized COVID-19 as a significant risk factor for the circulatory system disease mortality and reduced the unmeasured confounding effects.

**Conclusions:** The *PERR_proxy_* approach extends the applicability of the original PERR method to terminal event studies, offering a promising solution for addressing unmeasured confounding. Additionally, the DID regression framework provides a computationally efficient alternative for parameter estimation in PERR-based studies. However, careful consideration is still required in *PERR_proxy_* for proxy events selection and other underlying assumptions of the PERR method to ensure valid results.

## Introduction

Unmeasured confounding is very common in observational studies across different fields, potentially introducing bias to the effect estimates and effecting the validity of research findings. This challenge is crucial in medical and epidemiological research, where observational studies frequently rely on electronic health records (EHRs) and administrative databases. In medical observational studies, unmeasured confounders such as lifestyle characteristics, and genetic predisposition may generate inherent differences between exposed and unexposed groups prior to the exposure. These unmeasured confounders, which are rarely captured in EHRs, can influence both treatment selection and outcomes. Consequently, the final observed differences between the two groups reflect the combination of both exposure effect and pre-existing differences caused by unmeasured confounding. Therefore, it is important to reduce the unmeasured confounding effects when we evaluate the exposure effect.

Many statistical methods have been developed to deal with this problem. Among these, instrumental variable analysis (IVA) is widely used to control confounding when the confounders influence both treatment assignment and outcomes. IVA relies on the availability of a valid instrument, which is strongly correlated with the exposure but unrelated to the outcome except through the exposure. While IVA is a powerful tool, its performance depends heavily on the precision and validity of the chosen instrument^20,24,25^.

Another method to control unmeasured confounding is prior event rate ratio (PERR) adjustment method, which was first introduced by Weiner *et al.* in 2008^1^. PERR is a before-and-after method that provides simpler and more intuitive alternatives to IVA. Compared to IVA, before-and-after methods do not rely on the precision of the instrument variable. PERR estimates the exposure effect by comparing event rates between exposed and unexposed groups before and after exposure. By leveraging pre-exposure events as the baseline, PERR effectively reduces the unmeasured confounding effects. It has demonstrated good performance in observational studies utilizing EHR data. Subsequent research by Weiner and colleagues showed that when standard statistical analyses differed from randomized controlled trial (RCT) results due to unmeasured confounding, PERR was able to produce hazard ratios (HRs) that were more closely aligned with the RCT results^1–4^.

However, PERR relies on certain core assumptions. These include: no subjects accept exposure prior the post period, time-invariant unmeasured confounders effects, event outcomes do not affect the exposure, and study events are non-terminal^1^.

Several efforts have been made to extend the application of PERR and improve its accuracy. On the basis of standard PERR, an estimator called *PERR_ALT_* has been proposed by Yu *et al*^4^. Unlike the standard PERR, which employs separate Cox proportional hazards models for the prior and post periods, *PERR_ALT_* uses the paired Cox regression model to estimate the HRs for the two periods. The paired Cox regression model accounts for the within-subject correlation between the two periods, thereby improving the accuracy of the PERR estimates. In addition, *PERR_ALT_* directly models the change in HRs within two groups between the two periods by using paired Cox regression, reducing bias introduced by nonlinearity. Further advancements were made by Lin *et al*^6^, who extended *PERR_ALT_* using a pairwise Cox likelihood function. This approach introduced a computationally efficient method for estimating standard errors (SEs) and confidence intervals (CIs) of PERR estimates via the observed information matrix. However, this method depends on the asymptotic properties of maximum likelihood estimation (MLE), which require large sample sizes to ensure stability. For sparse data or complex models, this approach can become computationally unstable and lead to unreliable estimates. Despite these advancements, both the PERR and *PERR_ALT_* rely on the non-terminal event assumption, which restricts their use in terminal event studies due to the lack of prior events.

Violation of PERR assumptions can result in substantial bias in exposure effect estimates. Simulations have shown that violations—such as time-varying confounder effects, prior events affecting treatment, and mortality during the post-treatment period—can all introduce significant bias into PERR estimates. In some scenarios, PERR bias may exceed that of conventional hazard ratio estimates, becoming more pronounced as the extent of assumption violations increases^5^.

The non-terminal event assumption particularly restricts the applications of PERR in mortality outcomes, which are common and crucial endpoints in medical research. To address the limitations of PERR in terminal event studies, a method called post-treated event rate ratio (PTERR) has been proposed^19^. It defines the period from the beginning of treatment to when exposed group discontinued the treatment as “as-treated” period, and the period from the end of “as-treated” period to the end of the follow-up or death occurs as “post-treated” period. HRs for these two periods are calculated, and PTERR is then defined as *HR_As_*_−*Trt*_/*HR_Post_*_−*Trt*_. This approach provides us with a novel idea to explore the PERR application in terminal events, offering insights into exposure effects over time. However, the PTERR method assumes that the treatment effect stops immediately after the as-treated period ends, which is often impractical in clinical research. Many treatments that have long-term or even permanent effects after the treatment has been stopped, like lifestyle changes and surgical interventions. In addition, PTERR requires careful definition of the as-treated and post-treated periods and precise matching of exposed and unexposed subjects in order to define the as-treated period for unexposed group. Any errors in defining these periods or matching could bias the results.

Computational efficiency of parameters estimation is also an important consideration in PERR. In prior studies, the SEs and CIs of PERR estimates were estimated by the bootstrap method^1–4^, which is computationally intensive, especially when the sample size is large. The quick estimation based on the observed information matrix^6^ has limitations as we mentioned above. Therefore, we need more robust and efficient methods that align well with the PERR to do the parameter estimation for PERR estimates. One of them is the difference in difference (DID) method^7^. Similar to PERR, DID is also a before-and-after method, and is an analogous framework to PERR. PERR can be viewed as an extension of DID to time-to-event data^20^. A key advantage of DID is its foundation in regression modeling, which provides a more efficient and computationally faster alternative for estimating SEs and CIs compared to the bootstrap method. However, the use of DID regression for parameter estimation within the PERR framework has not been explicitly mentioned in previous research that focus on PERR. Additionally, most previous studies have applied DID regression to linear outcomes rather than time-to-event data modeled using Cox regression.

In this study, we propose a novel approach to extend the applicability of PERR in terminal event studies by introducing a proxy event to terminal event in the pre-exposure period. This proxy event framework explores the potential of the PERR applications in terminal event studies, providing a tool for reducing bias in exposure effect estimation. In addition, we aim to enhance the computational efficiency of parameter estimation for PERR estimates by integrating DID regression. Simulation studies are conducted under various scenarios to evaluate the method performance. We also conduct an empirical study to investigate the impact of severe hospitalized COVID-19 on circulatory system disease mortality by using our proposed methods.

## Methods

### 2.1 PERR method

The PERR method evaluates differences in event rates between exposed and unexposed groups during both the post-exposure and pre-exposure periods. Unlike conventional methods that focus solely on post-exposure differences, PERR accounts for baseline differences between the two groups prior to exposure. A key assumption of PERR is that no subjects in either the exposed or unexposed group accept exposure before the study period. Therefore, differences between the two groups observed before the exposure are entirely caused by the unmeasured confounding and are independent of the exposure effect. Given the presence of pre-exposure differences, the HR observed after the exposure (*HR_post_*) reflects both the effect of the exposure and the baseline differences due to unmeasured confounding between the groups. This makes *HR_post_* a biased estimator of the true exposure effect. PERR corrects for this bias by dividing *HR_post_* by the HR observed in the pre-exposure period (*HR_prior_*), as shown in (1):

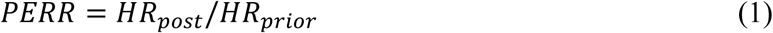

Alternatively, the formula above can also be calculated by using incidence rate ratio (IRR) instead of HR^1^.

### 2.2 Model Construction

The whole study period is divided into two distinct intervals:

- The *prior period* (*t*_1_): the time from the start of the study to the exposure.
- The *post period* (*t*_2_): the time from the exposure to the study endpoint.

The events of interest occurring in the two periods are called *prior event* (*y*_1_) and *post event* (*y*_2_), respectively. It is important to note that the start of *t*_2_ is not always appropriately defined as the time of receiving exposure; instead, it should correspond to the time when subjects first become at risk of experiencing *y*_2_^5^. However, for simplicity, we define the start of *t*_2_ as the time of receiving exposure in this study, and this definition will be used consistently in all subsequent analyses^5^.

If the event of interest is a terminal event (e.g., death), the data of *t*_2_ will only include survivors from *t*_1_, as those who experienced the terminal event during *t*_1_ have been dropped out (e.g., died). This causes the populations analyzed in the prior and post periods to become different, leading to severe selection bias in the PERR estimation. In addition, in general, exposure information is only available for subjects who survived the prior period, and the exposed and unexposed groups are not predefined at baseline in many cases, which will make the PERR infeasible since its denominator will be 0. Therefore, original PERR cannot be directly used in studies involving terminal events, we should develop approaches that can solve the selection bias and ensure valid PERR estimation.

In this study, we propose a novel approach that sets up a proxy event to substitute for the original terminal prior event, making the PERR become available in mortality event studies. To implement the new approach, we first identify a non-terminal event occurring in *t*_1_ that is highly correlated with the original terminal event. For example, in a study of COVID-19 mortality, hospitalization due to severe COVID-19 could serve as an appropriate proxy event. Then we exclude subjects who experienced terminal events during *t*_1_ and use proxy event as *y*_1_ to calculate HR in the remaining subjects, called *HR_proxy_*. The event definition and calculation of *HR_post_* in *t*_2_ remain unchanged. Thus, we can define a new “proxy” PERR estimate as follows:

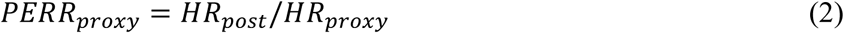

The schematic of *PERR_proxy_* is shown in **Figure 1**.

**Figure 1.**
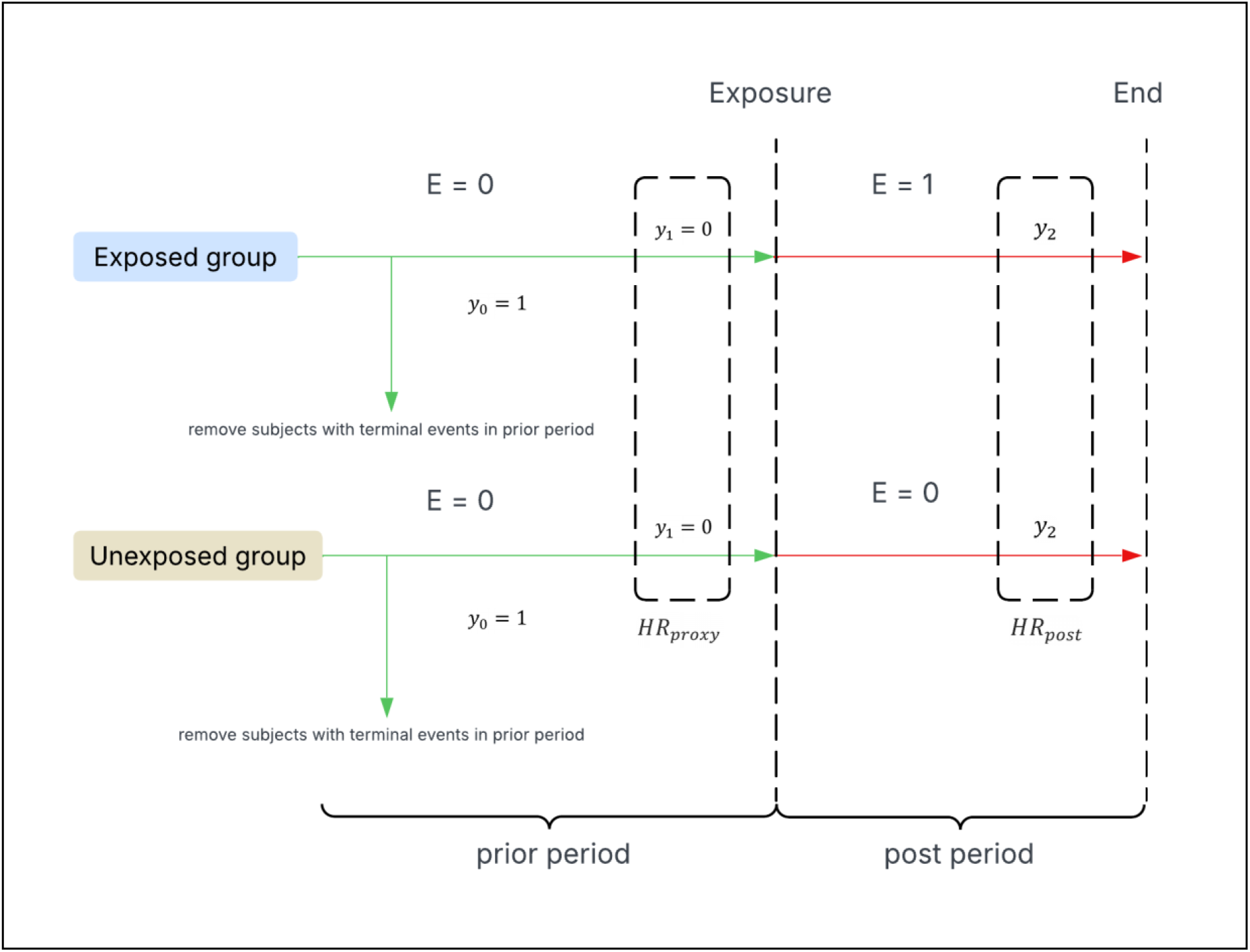
PERR_proxy_ method diagram. E denotes the exposure status of subjects. y_1_ and y_2_ denote the terminal outcome in the prior period and post period, respectively.

To improve computational efficiency in estimating SEs and CIs for the unmeasured confounding-adjusted estimates, we employed DID regression framework^6^. DID estimator for the *i*th subject can be calculated by using the following regression model:

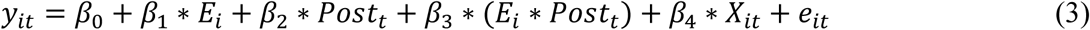

Where:

- *E_i_* is the exposure indicator (*E_i_* = 0 for no exposure and *E_i_* = 1 for exposure),
- *Post_t_* is the post period indicator (*Post_t_* = 0 for prior period and *Post_t_* = 1 for post period),
- *y_it_*, *X_it_* and *e_it_* represent the outcome, covariates and residual of the *i*th subject at time *t*, respectively.

This regression is fitted to the event of interest, and the estimated coefficient of the interaction term 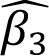 quantifies the relative change in *y_it_* from prior to post period in the exposed group relative to the unexposed group. DID regression can estimate the SEs and CIs through its regression framework directly, without relying on bootstrap methods as PERR does.

### 2.3 Simulation

#### 2.31 Simulation Objectives

In this simulation, we aim to:

i. Evaluate the performance of *PERR_proxy_* in correcting unmeasured confounding effects across different scenarios.
ii. Compare the SE and CIs estimation methods, with a focus on computational efficiency.

For aim (i), we evaluated the bias, mean squared error (MSE), CI coverage rates of *PERR_proxy_* estimates. For aim (ii), we assessed the computational time required for SEs and CIs estimation using different methods.

#### 2.32 Simulation set up

To explore these objectives, we constructed a series of scenarios by varying three key parameters:

a. Baseline risk,
b. Effect size of unmeasured confounding, and
c. Correlation between the proxy event and the terminal event.

We simulated time-to-event data incorporating both measured and unmeasured confounders, as well as scenario-specific covariates. In this simulation, we assumed that the unmeasured confounders (values and effects) remained constant across prior and post periods. For proxy events, we allowed the magnitude of unmeasured confounder effects to differ from those on the original terminal event, modulated through a pre-defined correlation parameter. This acknowledges that while proxy events can capture the same underlying risk structure of the original terminal event, they may express these effects differently. This specification maintains the time-invariance assumption of unmeasured confounders in the PERR method^1–4^ while accommodating the reality and characteristics of the proxy events.

The simulation included 10,000 subjects with a maximum follow-up time of 5 years. In each scenario, the simulation was repeated 200 times.

#### 2.33 Data generating mechanisms

The following notations were used to define the data generating process: *y_proxy_*, *y*_1_, *y*_2_ denote the proxy event (outcome), terminal prior event (outcome), and terminal post event (outcome), respectively. *X*_1_ and *X*_2_ denote measured confounders, while *UX*_1_ and *UX*_2_ denote unmeasured confounders. *E* denotes exposure indicator ( *E* = 0 for unexposed and *E* = 1 for exposed). (*Z*_1_,*Z*_2_), *p*, and *q* denote specific covariates in the exposure model, proxy outcome model, and prior outcome model respectively. *γ* denote the correlation between proxy event (y_proxy_) and terminal events (*y*_1_ and *y*_2_).

The measured and unmeasured confounders were generated as follows:

**Measured confounders:**

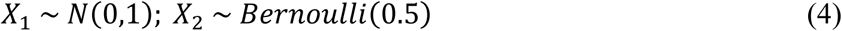

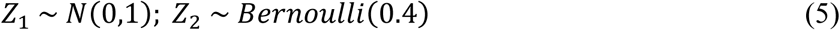

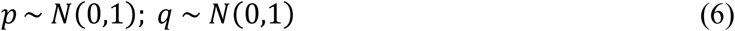

**Unmeasured confounders:**

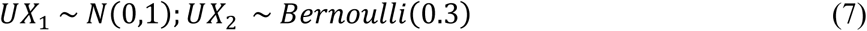

**Exposure Generation:**

The binary exposure variable *E* was generated using a log-linear model incorporating both measured and unmeasured confounders through (8):

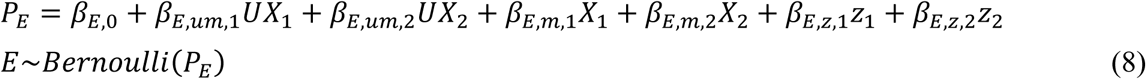

The parameter values for the exposure model were: *β*_*E*,0_ = 0; *β*_*E,um*,1_ = 1.2; *β*_*E,um*,2_ = 1.5; *β*_*E,m*,1_ = 0.1; *β*_*E,m*,2_ = *β*_*E,z*,1_*z*_1_ = *β*_*E,z*,2_*z*_2_ = 0.2.

**Outcome Generation:**

We simulated three time-to-event outcomes (*y_proxy_*, *y*_1_, *y*_2_) using hazard functions specified through linear predictors, incorporating relevant covariates. Survival times *T* for the outcomes (*y_proxy_*, *y*_1_, *y*_2_) were simulated using Weibull distribution with a fixed shape parameter *v* = 1.5 and a scale parameter *λ*(*x*) = *λexp* (*β^′^x*)^8^.

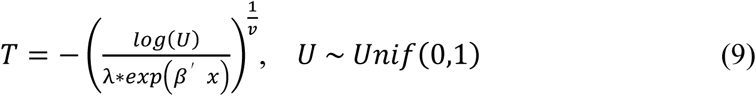

The hazard functions for the three outcomes were defined as:

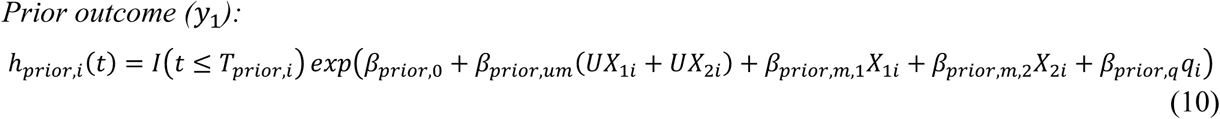

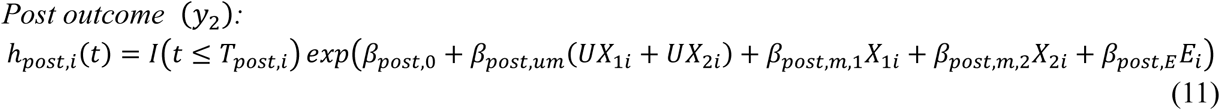

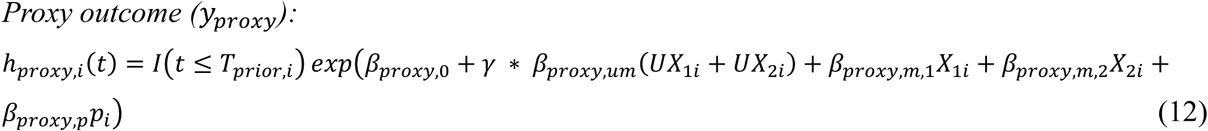

**Parameter specification:**

*β*_*prior*,*m*,1_ = *β*_*post,m*,1_ = *β*_*proxy*,*m*,1_ = 0.1; *β*_*prior*,*m*,2_ = *β*_*post*,*m*,2_ = *β*_*proxy*,*m*,2_ = 0.3; *β*_*prior*,*q*_ = *β*_*proxy*,*p*_ = 0.2; *β*_*post,E*_ = 0.5. *I*(*t* ≤ *T*_*prior,i*_) and *I*(*t* ≤ *T_post_*_,*i*_) were the indicators for the prior and post period respectively, both periods had a maximum censoring time of 5 years. *β*_*prior*,0_ = *β*_*post*,0_ = *β*_*proxy*,0_ varied between -5.3 to -2.3 to simulate different baseline risks. *β*_*prior,um*_ = *β*_*post,um*_ = *β*_*proxy,um*_ varied between 0.1 to 4.8 to simulate different unmeasured confounding effect sizes. *γ* varied between 0 to 1 to simulate different levels of correlation between *y_proxy_* and *y*_1_ (*y*_2_).

### 2.34 Data analysis

We fitted Cox proportional hazards regression to both *y*_2_ and *y_proxy_*, excluding subjects for whom *y*_1_ **=** 1 (i.e., those who experienced the terminal event during the prior period).

The *PERR_proxy_* was calculated by (2), the conventional HR was calculated by Cox proportional hazards regression model fitted to the post period data only. In this simulation, we have prior knowledge of subjects grouping information at the start of the study, as well as the mortality cases within each group. Therefore, we were able to calculate the standard PERR estimates. In order to assess sensitivity to non-terminal assumption violations, standard PERR estimates based on *y*_1_ and *y*_2_ models were calculated using equation (1), denoted as *PERR_original_*. The *DID_proxy_* estimates was calculated by *exp*(*β_interaction_*) where *β_interaction_* was the coefficient of the interaction term of period and exposure.

Bootstrap SEs and 95% CIs were computed using 1,000 iterations, with CIs derived from the 2.5th and 97.5th percentiles of the bootstrap estimates. The computational time for bootstrap and DID regression methods was recorded to evaluate efficiency.

For graphical comparisons of simulation performance, absolute bias was used to evaluate the magnitude of estimation error irrespective of direction. We also quantified the advantage of *PERR_proxy_* over each comparator as the comparator method’s absolute bias minus the absolute bias of *PERR_proxy_*, where positive values indicated better performance of *PERR_proxy_*.

All simulations and analyses were performed in R (version 4.3.2).

### 2.4 Application: the effect of severe COVID-19 on the circulatory system disease mortality

#### 2.41 Background

Understanding the relationship between severe COVID-19 and circulatory system diseases (CIR) mortality is critical due to the long-term sequelae of COVID-19. Common CIR diseases include ischemic heart disease, cerebrovascular disease, and hypertensive disorders, etc. Previous studies have identified COVID-19 as a potential risk factor for CIR diseases^9–14^, the virus of COVID-19 can attack the circulatory system by diverse biological mechanisms^15–17^. RCTs are rare in studying the COVID-19’s effect on CIR diseases due to the impracticality of randomizing COVID-19 exposure and the urgent nature of the pandemic, which needs rapid data collection instead of time-consuming trial designs. Therefore, most studies rely on observational studies, and the risk of suffering bias from unmeasured confounding increases.

To address these challenges, we applied the *PERR_proxy_* method to investigate severe COVID-19’s effects on CIR disease mortality. We implemented our approach using the UK Biobank (UKBB) data, a large-scale longitudinal cohort study comprising extensive health and genetic data from over 500,000 participants. Using the latest UKBB data, we extracted baseline characteristics and COVID-19 test records. Mortality events were obtained from national registry data. Cardiovascular system outcomes were defined using ICD-10 codes based on Clinical Classifications Software Refined (CCSR) categories.

#### 2.42 Model specification

In this study, we defined severe hospitalized COVID-19 as the exposure, CIR disease-related hospitalizations before the exposure as the proxy event, and CIR disease mortality after the exposure as the post event. After excluding subjects with error data (e.g., exposure after study end or proxy event before study start), 353,914 subjects were analyzed, 16,169 of them experienced severe hospitalized COVID-19, while 337,745 of them had no COVID-19 diagnosis. Potential measured confounders like sex, hypertension (HTN) history, coronary artery disease (CAD) history, and stroke history were incorporated into survival analysis. Nevertheless, other unmeasured confounders like genotypes and daily life habits remained unaccounted for.

The 6-month window was selected for both the prior and post periods. Given that the first COVID-19 case in the UK Biobank was documented on January 31, 2020, the prior period for all subjects was defined from August 3, 2019, to January 30, 2020, to minimize the influence of potential pre-existing COVID-19 exposure. This ensured that differences observed between the exposed and unexposed groups during the prior period were only caused by the patient’s baseline characteristics, independent of exposure effects. For the exposed group, the date of hospitalization due to severe COVID-19 was designated as the start of the post period. For the unexposed group, we used the prescription time-distribution matching method to assign hypothetical exposure dates based on the observed distribution of exposure dates in the exposed group^18^. While this definition of post period start point simplified our illustrative example, it is important to acknowledge that such methodology may be inappropriate when the exposure date does not align with the onset of post-event risk, as we mentioned previously^5^.

To account for competing risks, the endpoint of the post period was defined as the occurrence of death from non-CIR causes or the study end date for mortality assessment, whichever came first. This framework allowed for a more accurate estimation of CIR-specific mortality while handling censoring events appropriately.

Following the methodological framework described above, we now specify the statistical models used in this empirical study. Let ***X_i_*** denote the covariates matrix of the *i*th subject, and *E_i_* denote the exposure indicator (*E_i_* = 0 for unexposed and *E_i_* = 1 for exposed). Two Cox proportional hazards models were fitted for the prior (proxy) and post periods as follows:

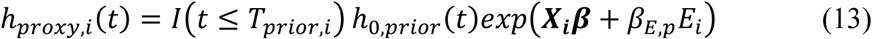

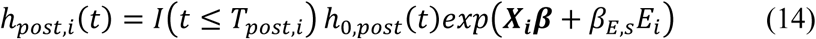

where *β_E_*_,*p*_ and *β_E_*_,*s*_ represented the coefficients of exposure in the prior and post periods respectively, ***β*** was the coefficient matrix of measured covariates.

The HRs for exposure in two periods were calculated as (15), and the *PERR_proxy_* estimate was then calculated by (2).

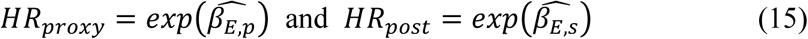

The corresponding 95% CI and SE of *PERR_proxy_* were estimated by bootstrap resampling with 1,000 iterations.

To contextualize our findings, we also calculated conventional HR for the post period without unmeasured confounding adjustment, along with the CIs, SEs, and p-value computational times for all approaches.

## Results

### 3.1 Simulation results^1^

#### 3.11 Performance of bias reduction and effect of β_0_, γ and intercept

Both *PERR_proxy_* and *DID_proxy_* showed very similar and much more substantial bias reduction ability compared to conventional HR and *PERR_original_*. The bias of *PERR_proxy_* and *DID_proxy_* generally decreased as *γ* increased, while *PERR_original_* and conventional HR maintained consistent bias across all values of *γ*. For example, when *β_um_* = 1.2 and intercept = -5.3, *PERR_proxy_* bias decreased from 2.5353 (*γ* = 0) to 0.0398 (*γ* = 0.9), and *DID_proxy_* bias decreased similarly from 2.516 (*γ* = 0) to 0.0433 (*γ* = 0.9). Conversely, the bias of *PERR_original_* and conventional HR maintained around -0.36 and 2.52, respectively regardless of *γ*. *PERR_original_* performed better than conventional HR and *PERR_proxy_* when *γ* was low.

Interestingly, *PERR_proxy_* exhibited negative bias as *γ* approached 1, indicating that its estimates tended to be biased toward the null, as our treatment effects were positive. When the unmeasured confounding effects were small (e.g. *β_um_* = 0.1) and the incidence of outcome was low (e.g. <10%), the magnitude of negative bias was small. However, when the unmeasured confounding effects and incidence increased, the negative bias became more prominent. Therefore, the absolute value of *PERR_proxy_* bias followed a U-shaped pattern with *γ* in such cases. *PERR_proxy_* bias was positive when *γ* was low, and the bias gradually decreased with increasing *γ*, then the bias turned negative and became more pronounced as *γ* approached 1. For example, when intercept = -2.3 and *β_um_* = 4.8, the bias of *PERR_proxy_* decreased from 1.2348 to 0.0732 as *γ* increased from 0 to 0.5; then the bias become negative and decreased from -0.0079 to -0.2093 as *γ* increased from 0.6 to 1. The absolute values of bias was the smallest under a moderately high level of *γ* (hereinafter referred to as γ*_optimal_*), but not when *γ* = 1. The γ*_optimal_* value systematically shifted depending on the strength of confounding (*β_um_*). For weaker confounding, γ*_optimal_* was approximately 0.8 – 0.9, while for stronger confounding, it shifted to 0.6 – 0.7. For example, when intercept = -4.7 and *β_um_* = 1.2, *γ_optimal_* = 0.9; when intercept = -4.7 and *β_um_* = 4.8, *γ_optimal_* = 0.7. This U-shaped relationship is summarized in **Figure 2**, which presents the absolute bias of *PERR_proxy_* across γ values under different intercept and *β_um_* settings. The figure shows that the lowest absolute bias was generally achieved at an intermediate-to-high value of *γ* rather than necessarily at *γ* = 1.

**Figure 2.**
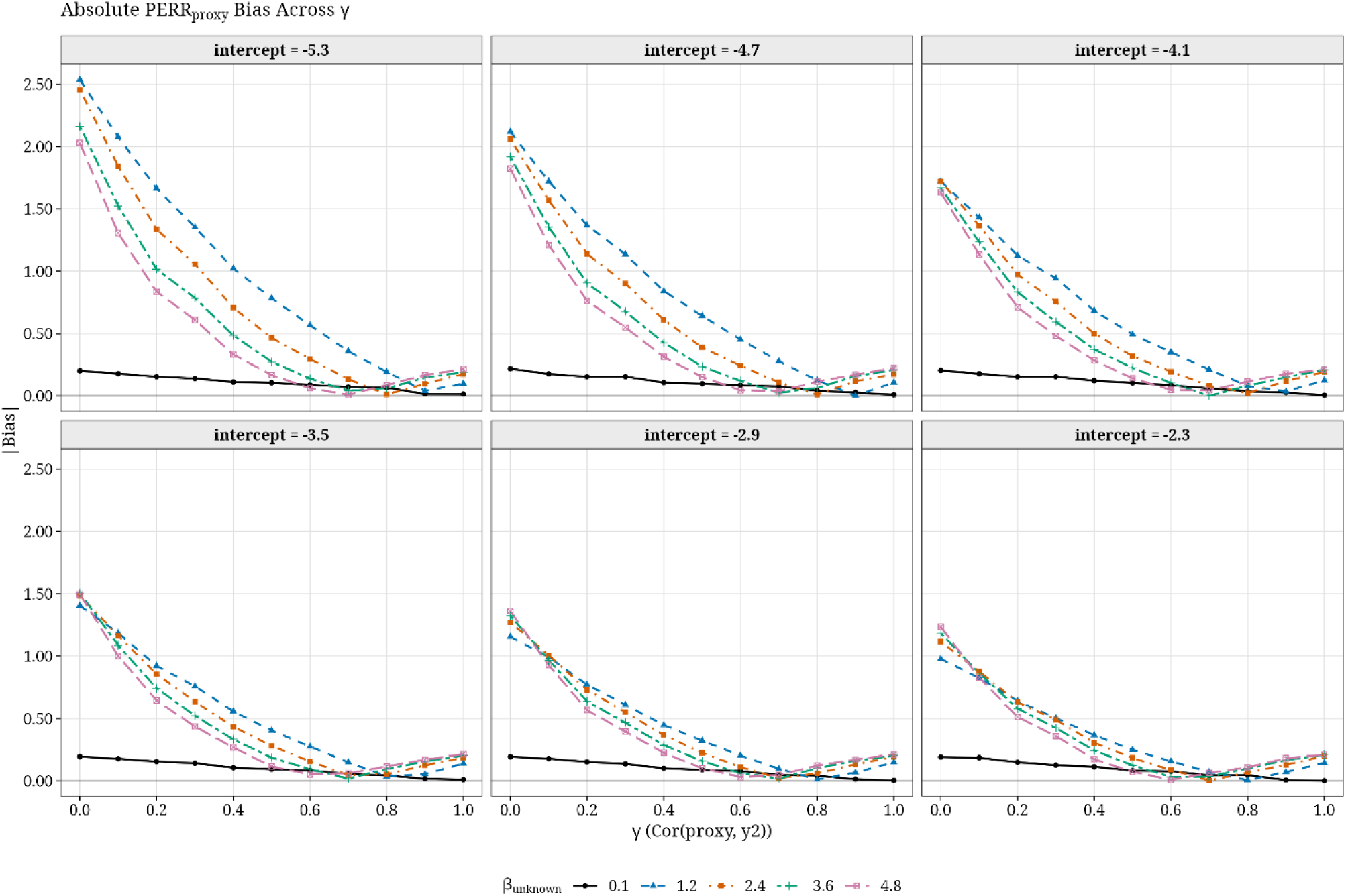
Absolute bias of PERR_proxy_ across different γ. Each panel corresponds to a intercept, and different curves represent strengths of unmeasured confounding, *β_um_*. The results show that the absolute bias generally decreased as γ increased, but in scenarios with stronger *β_um_*, and the *γ_optimal_* will change as intercept change.

The effects of unmeasured confounders (*β_um_*) and baseline risk (controlled by *β*_0_) significantly influenced method performance. Both conventional HR and *PERR_proxy_* showed decreasing bias as baseline HR increased. For example, when *γ* = 0.5 and *β_um_* = 1.2, conventional HR estimation, *PERR_proxy_* bias decreased from 2.5037 and 0.7828 (intercept = -5.3) to 0.9614 and 0.2468 (intercept = -2.3), respectively. In contrast, *PERR_original_* showed a slight increasing trend in bias (in absolute value), increasing from -0.3632 to -0.5451.

The effects of unmeasured confounding (*β_um_*) on performance of the methods was more complex. For conventional HR, *β_um_*’s effects on the performance showed a notable interaction with baseline risk (controlled by *β*_0_). At smaller intercepts (e.g., -5.3, -4.7), increasing *β_um_* reduced bias, while at larger intercepts (e.g., -2.9, -2.3), increasing *β_um_* increased bias. For example, when *γ* = 0.5 and intercept = -4.7, conventional HR bias decreased from 2.0768 (*β_um_* = 1.2) to 1.8006 (*β_um_* = 4.8), and when *γ* = 0.5 and intercept = -2.3, conventional HR bias increased from 0.9614 (*β_um_* = 1.2) to 1.2157 (*β_um_* = 4.8).

*PERR_original_* bias showed a consistent increasing trend with increased *β_um_* under all intercepts, except in scenarios with negligible confounding (*β_um_* = 0.1). For example, when *γ* = 0.5 and intercept -4.7, its bias (in terms of absolute value) increased from -0.4163 (*β_um_* = 1.2) to -0.7648 (*β_um_* = 4.8). Similarly, when *γ* = 0.5 and intercept -2.3, its bias (in absolute value) increased from -0.5451 (*β_um_* = 1.2) to -0.7959 (*β_um_* = 4.8).

*PERR_proxy_* bias exhibited distinct trends depending on whether *γ* was below or above *γ_optimal_*. Specifically, for *γ* ≤ *γ_optimal_*, increasing *β_um_* tended to reduce bias. For *γ* ≥ *γ_optimal_*, bias either decreased and then increased or consistently increased. For example, when intercept was -5.3, *γ_optimal_* were between 0.7 and 1 under different *β_um_*. When *γ* = 0.2, *PERR_proxy_* bias decreased from 1.6639 (*β_um_* = 1.2) to 0.8356 (*β_um_* = 4.8); when *γ* = 0.8, *PERR_proxy_* bias decreased from 0.1919 (*β_um_* = 1.2) to 0.0121 (*β_um_* = 2.4) first, then increased to -0.0869 (*β_um_* = 4.8) (in absolute value); when *γ* = 0.9, *PERR_proxy_* bias increased from 0.0398 (*β_um_* = 1.2) to -0.1676 (*β_um_* = 4.8) (in absolute value). This trend confirmed that *γ_optimal_* served as a critical threshold for optimal performance in the relationship between *PERR_proxy_* bias and *β_um_*. In addition, the bias of conventional HR and *PERR_original_* remained relatively stable across different values of *γ* as they did not incorporate with proxy event framework. The overall bias advantage of *PERR_proxy_* over the other methods is shown in **Figure 3**. Positive values indicate scenarios in which *PERR_proxy_* achieved smaller absolute bias than the comparator. The advantage was most evident when compared with conventional HR and *PERR_original_*, whereas the difference between *PERR_proxy_* and *DID_proxy_* was generally small, consistent with their similar bias patterns.

**Figure 3.**
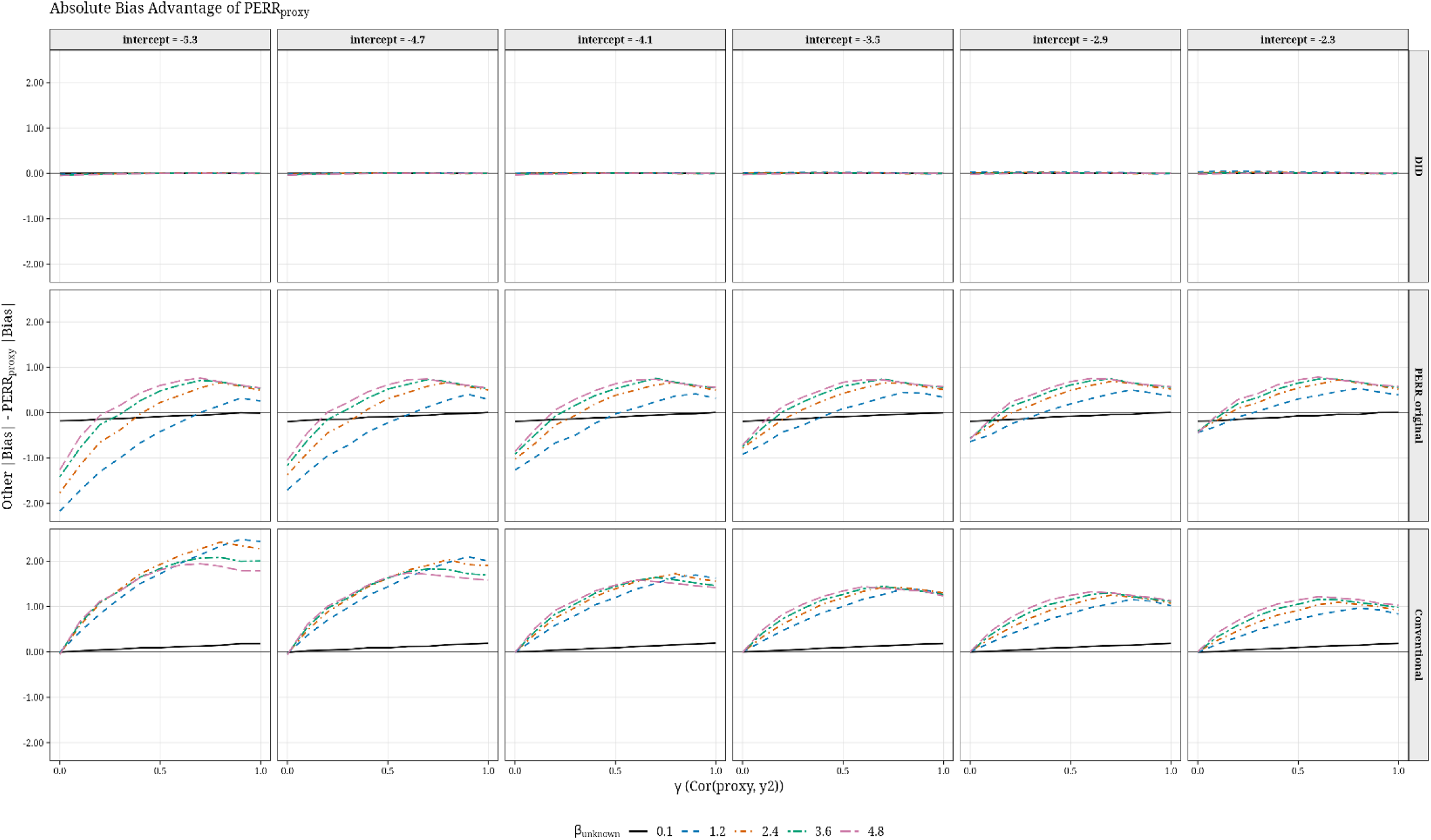
Absolute bias advantage of PERR_proxy_ over comparator methods. Across most scenarios, *PERR_proxy_* showed clear advantage compared with conventional HR and *PERR_original_*, particularly when *γ* was moderate to high. Its performance was closely aligned with *DID_proxy_*, reflecting the conceptual similarity between the two approaches.

Additional visual summaries of bias, MSE, and relative improvement across the full simulation are provided in Supplementary **Figures S1–S5**.

#### 3.12 Standard error estimation and computational efficiency

We evaluated two approaches for SE estimation: bootstrap and DID regression. The estimated SEs by DID regression were generally lower than those of bootstrap across different scenarios. For example, when intercept = -4.7 and *β_um_* = 2.4, the SEs given by DID regression were around 0.1 while the SEs given by bootstrap were from around 0.12 to 0.39.

Despite differences in SEs, both methods produced similar CI coverage rates for *PERR_proxy_*. Both methods showed a U-shaped relationship between CI coverage and *γ* like the bias we observed in Section 3.11. For example, when intercept = -4.1 and *β_um_* = 3.6, the highest CI coverage rate was achieved at *γ* = 0.8, with coverage rate decreased as *γ* continually increased toward 1.

Across all scenarios, the DID regression consistently showed superior computational efficiency compared to the bootstrap in SEs and CIs estimation. For example, when intercept = -4.7, *β_um_* = 2.4, and γ = 0.5, DID regression required ∼0.2 seconds to compute the estimate SE and CI, while the bootstrap (with 1,000 iterations) required ∼6.8 seconds. DID regression was about 41 times faster than the bootstrap in this case.

#### 3.2 Empirical results

**Table I** summarizes the results of the empirical analysis, including estimates for *PERR_proxy_*, *DID_proxy_*, conventional HR, and HR for the prior period (*HR_prior_*), along with their 95% CIs, SEs, p-values, and computational times.

**Table I.**
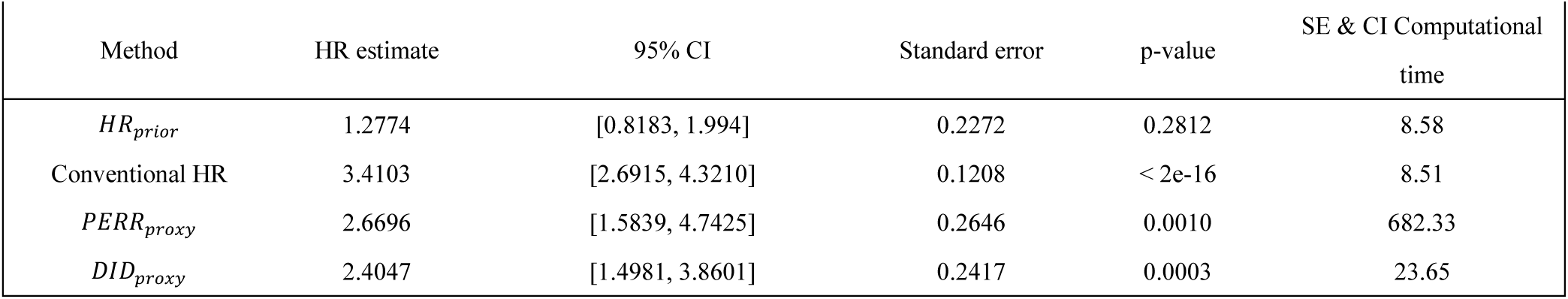
Comparative Analysis of Hazard Ratio Estimates for Effect of Severe Hospitalized COVID-19 on Circulatory Disease Mortality.

Both *PERR_proxy_*, *DID_proxy_* and conventional HR estimates were significantly greater than 1 with (p-value < 0.05), indicating that severe hospitalized COVID-19 is a significant risk factor to the CIR disease mortality. However, the conventional HR yielded the highest HR estimate, which was notably larger than the adjusted estimates from *PERR_proxy_* and *DID_proxy_*. This suggests that the conventional HR overestimated the exposure effect due to its inability to account for unmeasured confounding.

The *PERR_proxy_* estimate and *DID_proxy_* estimate were closely aligned, reflecting the high conceptual similarity between the two methods. Additionally, *HR_prior_* was close to 1 and not statistically significant (p-value = 0.28122), suggesting that there were no significant baseline differences between the exposed and unexposed groups prior to the exposure.

Finally, in terms of computational efficiency, DID regression demonstrated substantial advantages over the bootstrap method for calculating CI and SE. This makes DID regression a more practical and efficient parameter estimating approach for large-scale analyses.

## Discussion and Conclusion

In this paper, we introduced a novel approach that expands the applicability of the PERR method to terminal events studies by incorporating a proxy event framework under appropriate assumptions. The numeric simulations and empirical study results demonstrated the great ability of this approach in reducing unmeasured confounding effects when evaluating the true effects of exposure. This advancement addresses a key limitation of the original PERR method mentioned by Weiner and colleagues^1–4^ that PERR cannot deal with terminal events or the events lacking prior events. In addition to presenting the proxy event extension of PERR, we also studied the parameter estimation methods, including the use of DID regression for efficient computation of SEs and CIs.

One of the key findings in the simulation was the U-shaped relationship between *γ* and *PERR_proxy_* bias. This finding challenged the intuitive belief that the proxy event should always be similar to the original terminal event as much as possible. Instead, our results suggest that some degree of difference between them is necessary to minimize bias. This highlights the importance of evaluating and selecting proxy events carefully.

We speculate that one possible reason underlying this U-shaped phenomenon is the opposite effects from unmeasured confounding and non-collapsibility of HRs in Cox regression. Non-collapsibility is an inherent property of nonlinear effect measures, such as HRs in Cox regression^21–23^, it causes the marginal HR (in which covariates are omitted) differ from the conditional HR (in which covariates are included). When covariates are omitted, the marginal HR becomes an averaged effect across strata of the omitted covariates. This averaging process compresses the HR toward 1 (the null), leading to an underestimation of the true exposure effect, and the magnitude depends on several factors including the strength of the exposure effect, the covariates effects and the censoring rates^22^. The non-collapsibility persists even in the absence of unmeasured confounding, making it an unavoidable source of bias in Cox regression^21^.

In our simulations, *HR_post_* represented the exposure effect in the post-period, which shifted towards the null due to non-collapsibility owing to omitted unmeasured covariates. However, the true causal exposure effect in the prior period is zero, hence it is not affected by non-collapsibility^22^. As such, *PERR_proxy_* (*HR_post_* / *HR_proxy_*) is shifted towards the null, creating a negative bias.

On the other hand, unmeasured confounding introduces bias in the opposite direction. When *γ* was low, unmeasured confounders are inadequately accounted for in the proxy event model. Therefore, a large part of unmeasured confounding effects were not removed after dividing *HR_proxy_*. Since the unmeasured confounders were positively associated with the outcome, the exposure effect estimate was inflated (biased away from the null) due to the inclusion of unmeasured confounding effects. However, as *γ* increased, more unmeasured confounding effects were captured by the proxy event framework, thereby reducing confounding bias. Beyond a certain point γ*_optimal_*, non-collapsibility in *HR_post_* became the dominant source of bias, so the observed *PERR_proxy_* shifted below the true exposure effect. Since non-collapsibility bias is more prominent when the covariates effects are strong^22,23^, the point at which non-collapsibility becomes the dominant source of bias occurs earlier under larger values of *β_um_*. This means the *γ_optimal_* values are smaller under large *β_um_*. This is consistent with the shifting pattern of γ*_optimal_* observed in our simulations. When the *γ* = 1, the remaining negative bias in the observed *PERR_proxy_* entirely reflects non-collapsibility, which systematically underestimates the exposure effect. This acknowledges that non-collapsibility is an intrinsic property of the Cox and cannot be eliminated, even without unmeasured confounding^21^.

The proposed *PERR_proxy_* builds on several core assumptions of the original PERR framework introduced by Weiner and colleagues^1–4^. These assumptions include (i) prior events do not affect the probability of exposure, (ii) the unmeasured confounding effect remains constant across two periods, (iii) no exposure prior to the post period. These assumptions must be carefully evaluated before applying *PERR_proxy_* to ensure its validity.

Additionally, the selection of an appropriate proxy event is critical. An appropriate proxy event should be a non-terminal event that is correlated with the original terminal event, so that can capture sufficient unmeasured confounding information while remaining some variability to offset part of the non-collapsibility effects. In real applications, the U-shaped relationship between *PERR_proxy_* bias and *γ* may offer some practical advantages as it is often impossible to find a perfect proxy event be identical and perfectly same to the original terminal event. The correlation coefficient *γ* is unlikely to be very high due to differences in the nature of the proxy and prior events, measurement error, and incomplete data. Instead, it is often moderate, which could close to the range of γ*_optimal_*. This suggests that the framework is suitable to handle realistic scenarios where proxy events are imperfect but still informative.

In conclusion, *PERR_proxy_* provides a new approach to reduce the unmeasured confounding effects in terminal event studies, building on the standard PERR framework. The incorporation of DID regression further enhances the method by providing computationally efficient estimates of SEs and CIs, making it suitable for large-scale datasets.

In future, the proxy-based framework can be adapted to the *PERR_ALT_* estimator proposed by Yu *et.al*^4^, which used the pairwise Cox regression model and could further reduce the bias caused by nonlinearity.

In addition, currently there are no clear criteria for proxy event selection, selection is made empirically. As demonstrated above, proxy event selection is crucial for *PERR_proxy_* as it requires an appropriate correlation with the original terminal events. Besides selecting a suitable proxy event, *PERR_proxy_* still relies on other assumptions of standard PERR. Future work could focus on developing quantitative criteria for proxy event selection and methods to evaluate the core assumptions underlying PERR. Furthermore, it would be valuable to investigate the impact of assumption violations on the performance of *PERR_proxy_*, and whether certain assumptions can be relaxed to further expand the applicability of *PERR_proxy_*.

## Supporting information

Simultion results

## Data Availability

The UK Biobank data is available to all registered researchers upon application. All other data produced in the present work are contained in the manuscript.

## Footnotes

1 The full simulation results are available at: https://mycuhk-my.sharepoint.com/:x:/g/personal/1155124429_link_cuhk_edu_hk/EeXDlxSle55Evz486U_kn3oBEq9a33k9euWwxZxGRO-xMA?e=SHCtmX

