## Supplementary Figure 1-5 for "A new approach using proxy event in prior event rate ratio for terminal event studies"

**Figure S1. Heatmap of absolute $PERR_{proxy}$ bias across simulation scenarios.** The heatmap provides an overview of the $PERR_{proxy}$ performance. Lighter cells indicate smaller absolute bias.

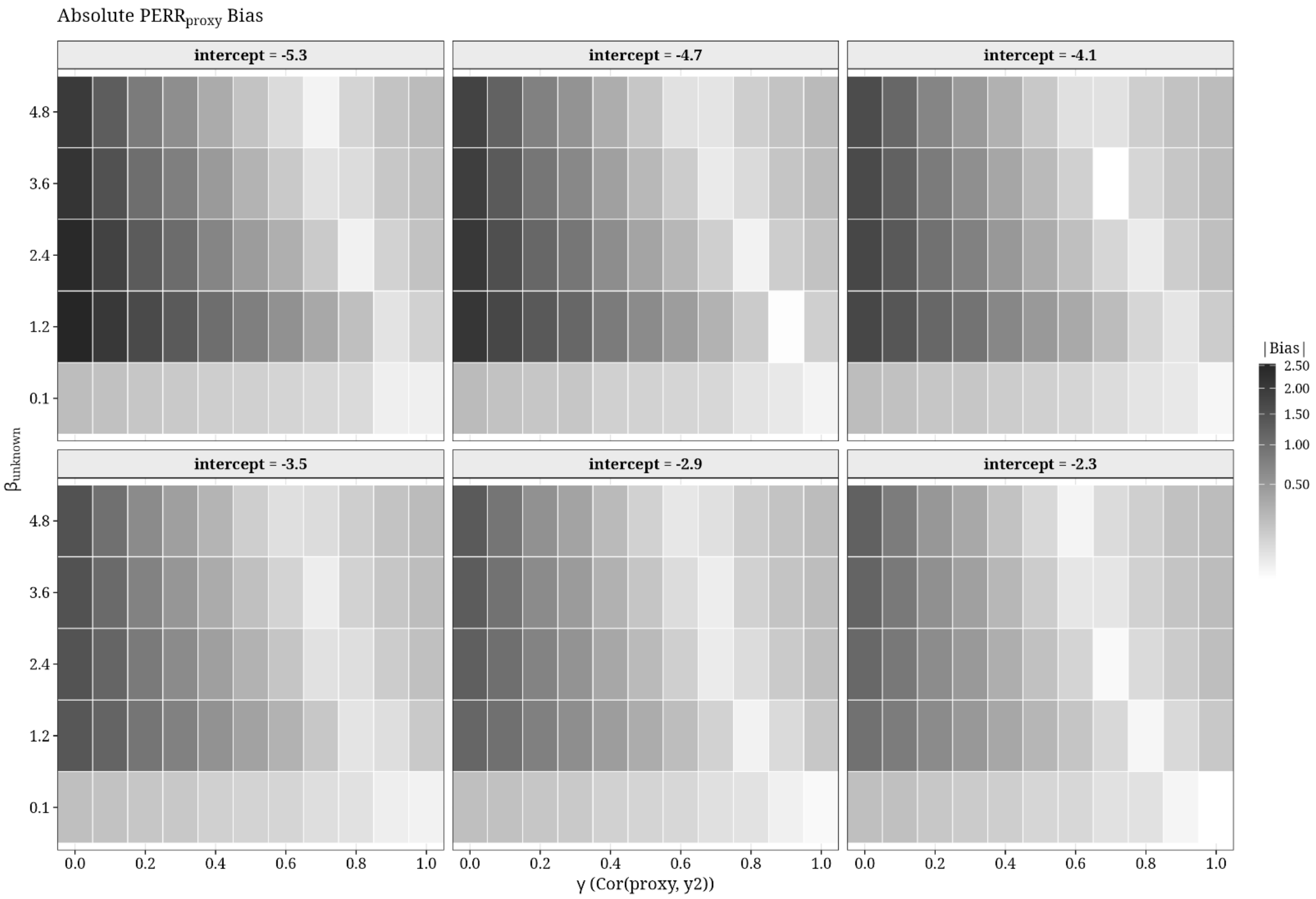

**Figure S2. Absolute bias comparison across methods.** Curves show the absolute bias for $PERR_{proxy}$, $DID_{proxy}$, $PERR_{original}$, and conventional HR across different $\gamma$ values, intercepts and $\beta_{um}$.

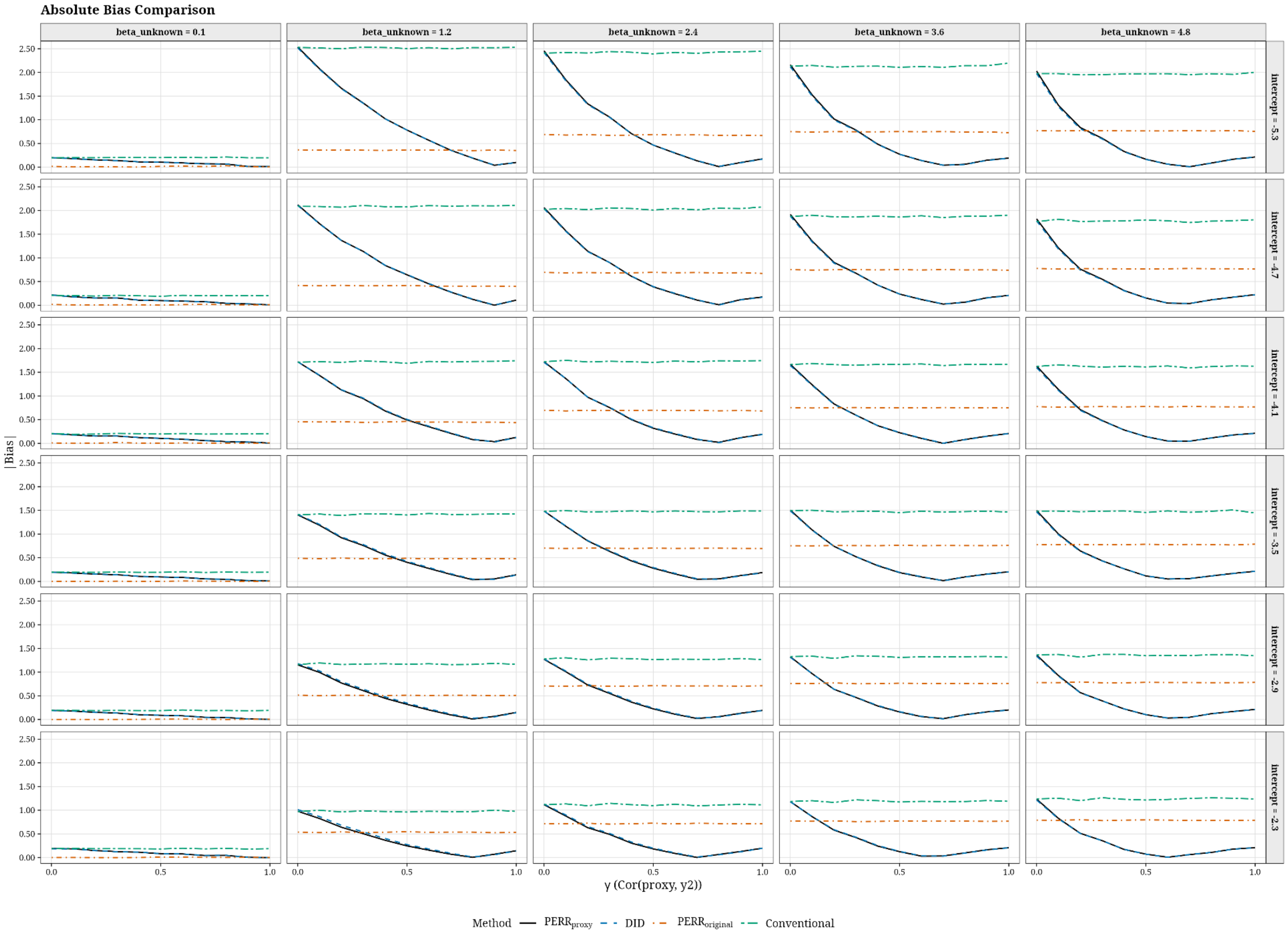

**Figure S3. MSE comparison across methods.** Curves show the MSE for $PERR_{proxy}$, $DID_{proxy}$, $PERR_{original}$, and conventional HR across different $\gamma$ values, intercepts and $\beta_{um}$.

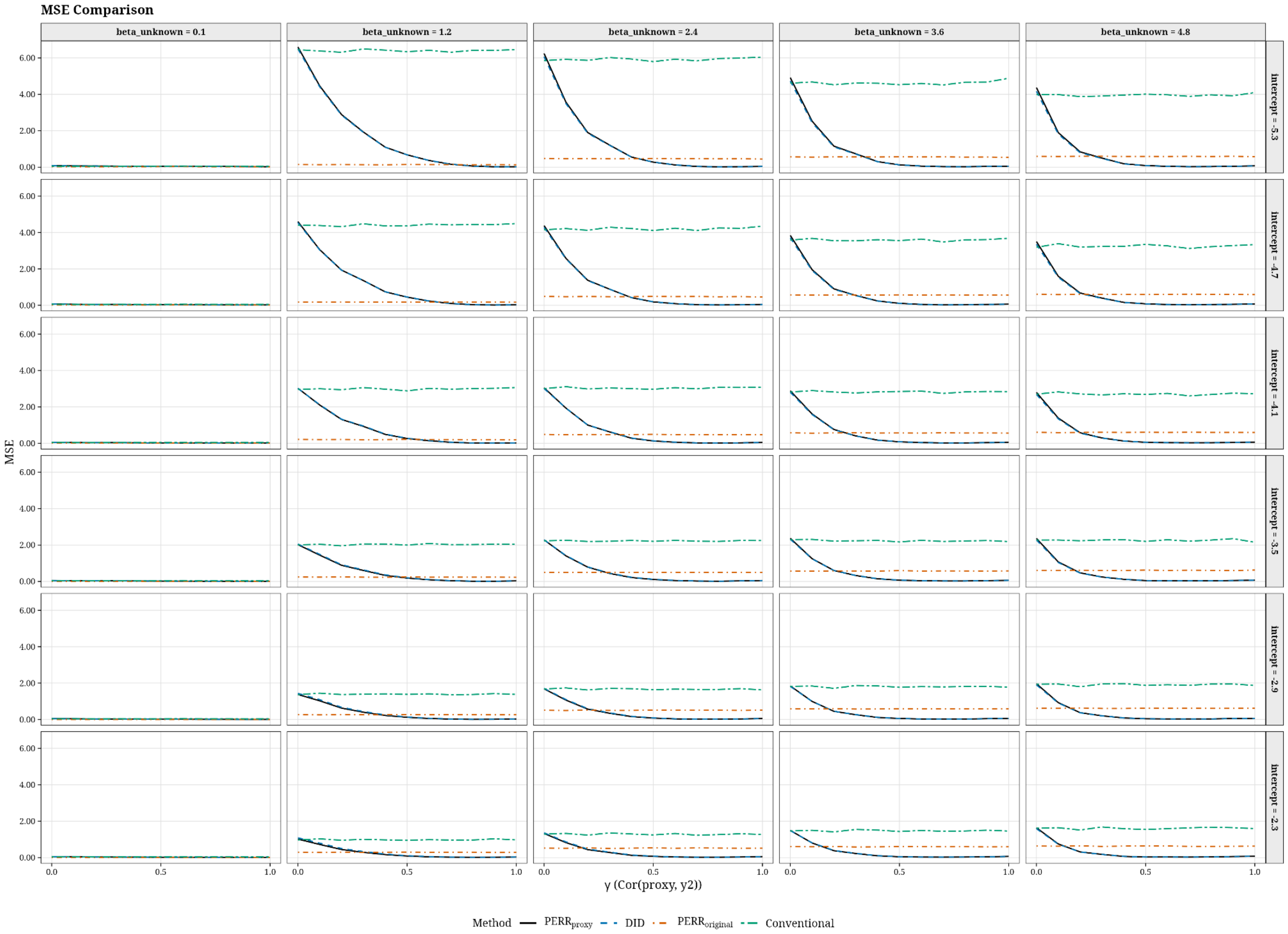

**Figure 4. MSE advantage of $\mathbf{PERR_{proxy}}$ over comparator methods**. Across most scenarios, $PERR_{proxy}$ showed clear advantage compared with conventional HR and $PERR_{original}$, particularly when $\gamma$ was moderate to high. Its performance was closely aligned with $DID_{proxy}$, reflecting the conceptual similarity between the two approaches.

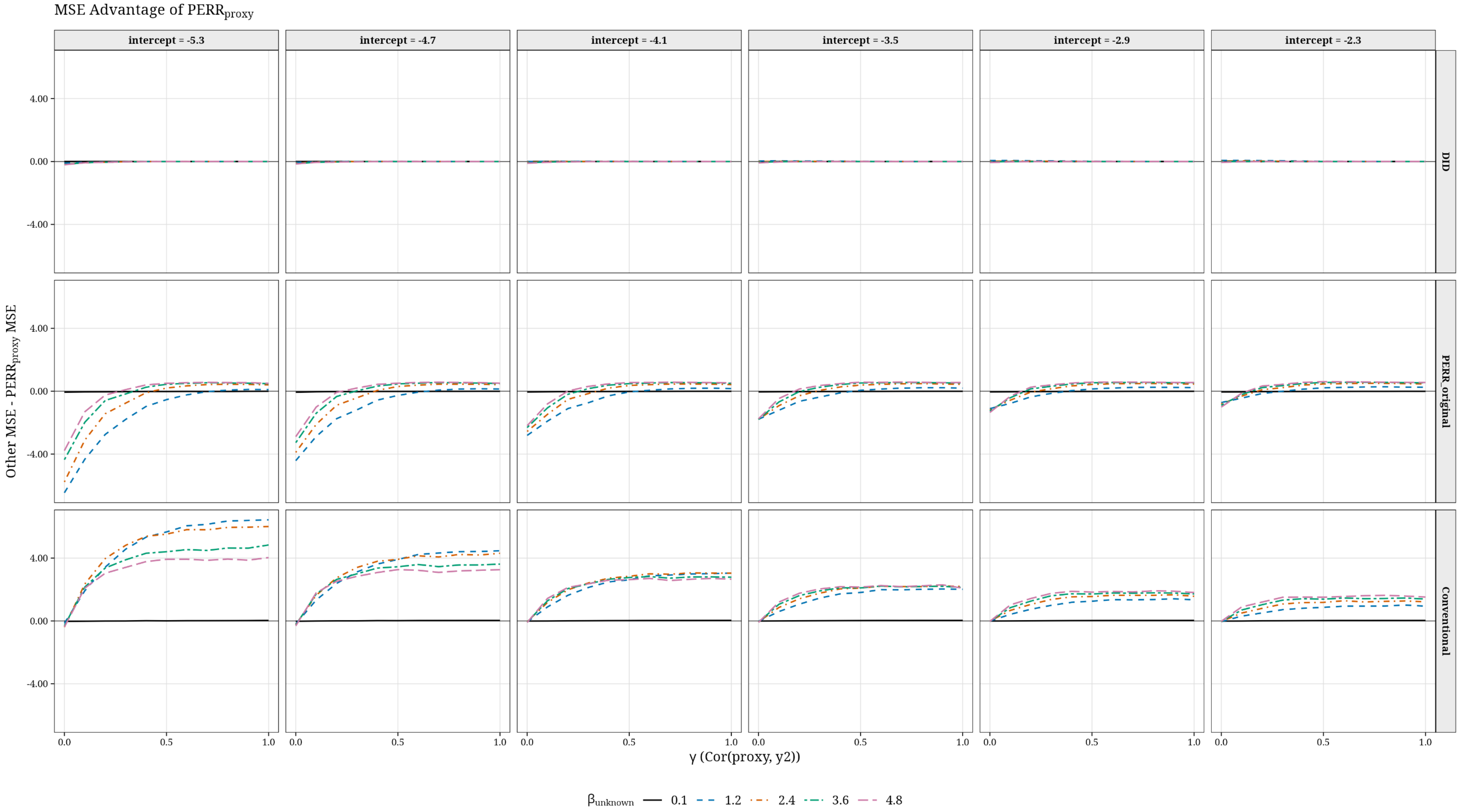

**Figure S5. Relative improvement in bias reduction of $PERR_{proxy}$ compared with conventional HR across $\gamma$.** Relative improvement was calculated as (conventional HR bias − $PERR_{proxy}$ bias) / conventional HR bias. It represents the proportional reduction in bias achieved by the $PERR_{proxy}$ approach relative to conventional HR approach. Higher values indicate greater bias reduction by $PERR_{proxy}$ across different $\gamma$ values, intercepts, and $\beta_{um}$.

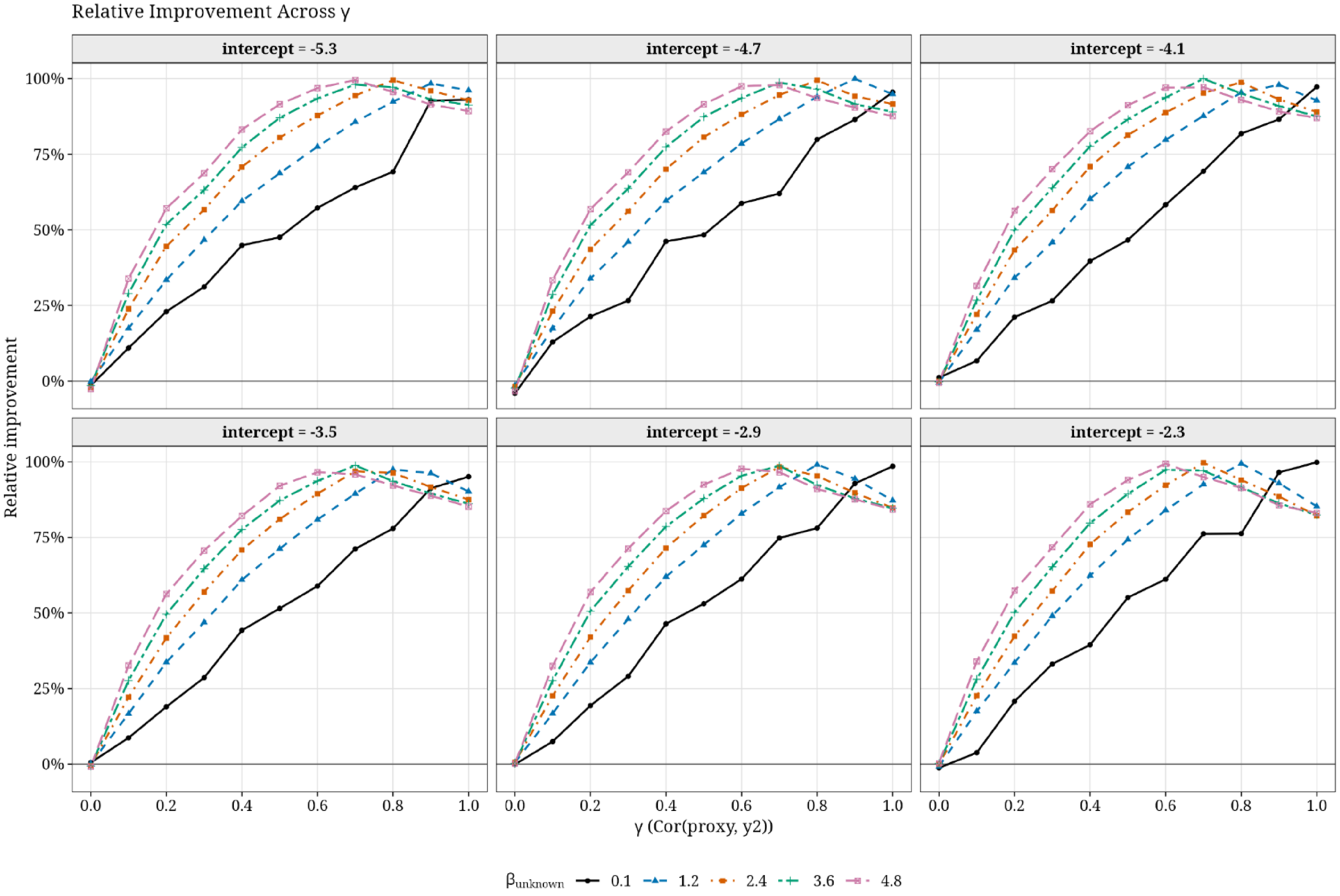
